# Accounting for comorbidity in etiological research

**DOI:** 10.1101/2025.01.19.25320775

**Authors:** Vahe Khachadourian, Magdalena Janecka

## Abstract

**Introduction:** Despite the theoretical advancements and recommendations regarding covariate adjustment in causal inference, clinical studies often fail to explicitly state the underlying assumptions related to causal structure among the study variables. Specifically, despite the pervasive nature of comorbidity, explicit causal assumptions about the role of comorbidity in exposure-outcome relationships are often lacking, potentially leading to inappropriate accounting for comorbid conditions and resulting in biased effect estimates. This study aims to explore common causal structures involving comorbidity and provide guidance for handling it in etiologic research.

**Methods:** We use Directed Acyclic Graphs (DAGs) to depict six causal scenarios involving comorbidity as a confounder, mediator, collider, or consequence of the exposure or outcome. Simulations were conducted across 5,000 iterations for each scenario, assessing the impact of conditioning on comorbidity under three effect measures (mean difference, odds ratio, risk ratio). Bias was evaluated by comparing adjusted and unadjusted effect estimates to the true values.

**Results:** The impact of conditioning on comorbidity varied by its causal role. Adjusting for comorbidity mitigated bias when it acted as a confounder, but introduced bias when it was a mediator or collider. In instances where comorbidity was a consequence of either the exposure or outcome, the decision to adjust depended on the research objectives. Nonlinear models revealed differences in marginal and conditional effects due to non-collapsibility.

**Discussion:** Explicit causal assumptions are essential for selecting appropriate analytical strategies in etiologic research. This study provides practical guidance on handling comorbidity-related challenges, highlighting the need for study design and analysis to align with research objectives. Future work should address more complex causal structures and other methodological challenges.

## INTRODUCTION

Over the past decades, etiologic research on human disease has advanced significantly due to improvements in causal inference methodology, availability of health records – including both registry-based studies and real-world data – and establishment of international consortia allowing to gauge the generalizability of results. Throughout, a considerable focus in observational studies has been placed on reducing confounding and other biases^1^. Although numerous publications ^2–5^ have now addressed various aspects of “adjustment” in design or analyses, many of these works are either theory-based or focused on specific causal structures^3,4^, limiting their wider implementation in applied research. Specifically, the issue of comorbidity, its various causal relationships with the outcome of interest, and downstream analytical recommendations have not been widely discussed. Therefore, addressing the issue of comorbidity in etiological studies, integrating clinical perspective and focusing on clinical concepts, could help bridge the gap between methodological knowledge and practice.

Comorbidity, first defined by Feinstein (1970)^6^, is the presence (existence or occurrence) of another distinct health condition in the individual with the index disease. Although there is variation in the definition and interpretation of comorbidity^7,8^, studies consistently show that comorbidity is a common phenomenon^9^. Given the pervasive nature of comorbidity in many disorders^10^, etiologic studies often consider comorbidity in their design or analyses^11–16^. The motivations for such considerations are different and could include understanding of the risk factors for the disease of interest “independent” of its comorbidity^17^, control of confounding^18^, minimizing misclassification^19^, or study effect measure modification by comorbidity^20^. Despite these various contexts for considering comorbidity in etiologic research, the underlying motivation is most often to reduce bias or obtain a valid estimate for the parameters of interest.

Given that there is no universally assumed (or universally true) causal structure between comorbidity^8^ and other variables in a study, a one-size-fits-all solution for accounting for comorbidity in etiologic research does not exist. For instance, comorbidity could be a risk factor for the index disease, its consequence, or share a common cause with it. Depending on the causal structure at hand, adjusting for comorbidity can change the effect estimate of the exposure on the outcome, sometimes resulting in attenuation, amplification, or even reversal of the estimated association relative to the comorbidity unadjusted associations^2–4,21^. Understanding the scenarios under which comorbidity adjustment is made and its impact on the main effect of exposure on the index diagnosis is therefore critical.

As the definition of comorbidity does not imply any explicit or implicit causal relation or even temporal order between the index disease and the co-occurring comorbid conditions, it is important to be clear about the assumed structure(s) – which often is critical in informing the correct design or analytical strategy. However, a review of the literature reveals that the handling of comorbidity in the design and analysis of etiological studies often lacks explicit assumptions about their relationship with other variables, or may in fact not align with the study objective^11,16^.

In this paper, we therefore explore several common causal structures involving comorbidity and other study variables. For each scenario, we discuss the impact of adjusting for comorbidity on the effect estimates and potential considerations. We hope this piece can provide guidance for accounting for comorbidity (and potentially other covariates) in epidemiological research and improve causal inference.

Controlling for comorbidity extends beyond adjustment in the analytical model and can occur by restricting the sample based on comorbidity, stratified sampling or analysis based on comorbidity, or handling differential loss to follow-up or non-random missingness patterns. Here forth, for ease of read we use conditioning and adjusting interchangeably, and unless otherwise specified, the arguments and statements about adjustment are applicable to all other conditioning approaches.

## METHODS

### Possible Structure of Associations

Explicitly stating the causal assumptions is critical for the choice of the study design or analytical methods, and clear communication and justification of these decisions. Causal diagrams, specifically Directed Acyclic Graphs (DAGs)^22^ are a common tool in causal inference for depicting assumptions and assessing bias. Concise and accessible description of the causal diagrams are provided by Greenland et. al (1999)^23^.

Here we use DAGs to depict several possible common scenarios underpinning the associations between comorbid conditions and the exposure and outcome in study. We use X to denote the key exposure, Y for the index condition, and C for the comorbidity.

1. Comorbidity as a common cause of X and Y
2. Comorbidity as a mediator of the effects of X on Y
3. Comorbidity as a collider in the association between X and Y
4. Comorbidity as a consequence of X, but not a risk factor for Y
5. Comorbidity as a risk factor of Y, but independent of X
6. Comorbidity as a consequence of Y, but not directly associated with X

### Effect measures

The impact of adjusting for comorbidity on the estimate of the total effect depends on the effect measures. To extend the applications of our work, for each of the above scenarios presented, we will assess and discuss the effect of conditioning on comorbidity for three different effect measures; 1) mean difference (least squares regression), 2) odds ratio (logistic regression), and 3) risk ratio (log binomial regression).

### Simulation setting

We conducted a simulation study to assess the effect of conditioning on covariate C in each of the causal structures presented in Figure 1. The parameters for the simulation analyses, along with the analytical code, are presented in the supplemental material. Briefly, for each of the causal structures presented in Figure 1, we made 5,000 iterations of simulated studies with sample size 1000. For simplicity, we assumed X and C are both binary variables. For each causal diagram we simulated 3 sets of outcomes (Y): two binary endpoints (e.g., hip fracture) corresponding to the total effect of X on Y on the RR and OR scales to be 2; and one normally distributed, continuous endpoint (e.g., serum cholesterol level) for which the total effect of X on Y measured by the mean difference was equal to 2.

**Figure 1.**
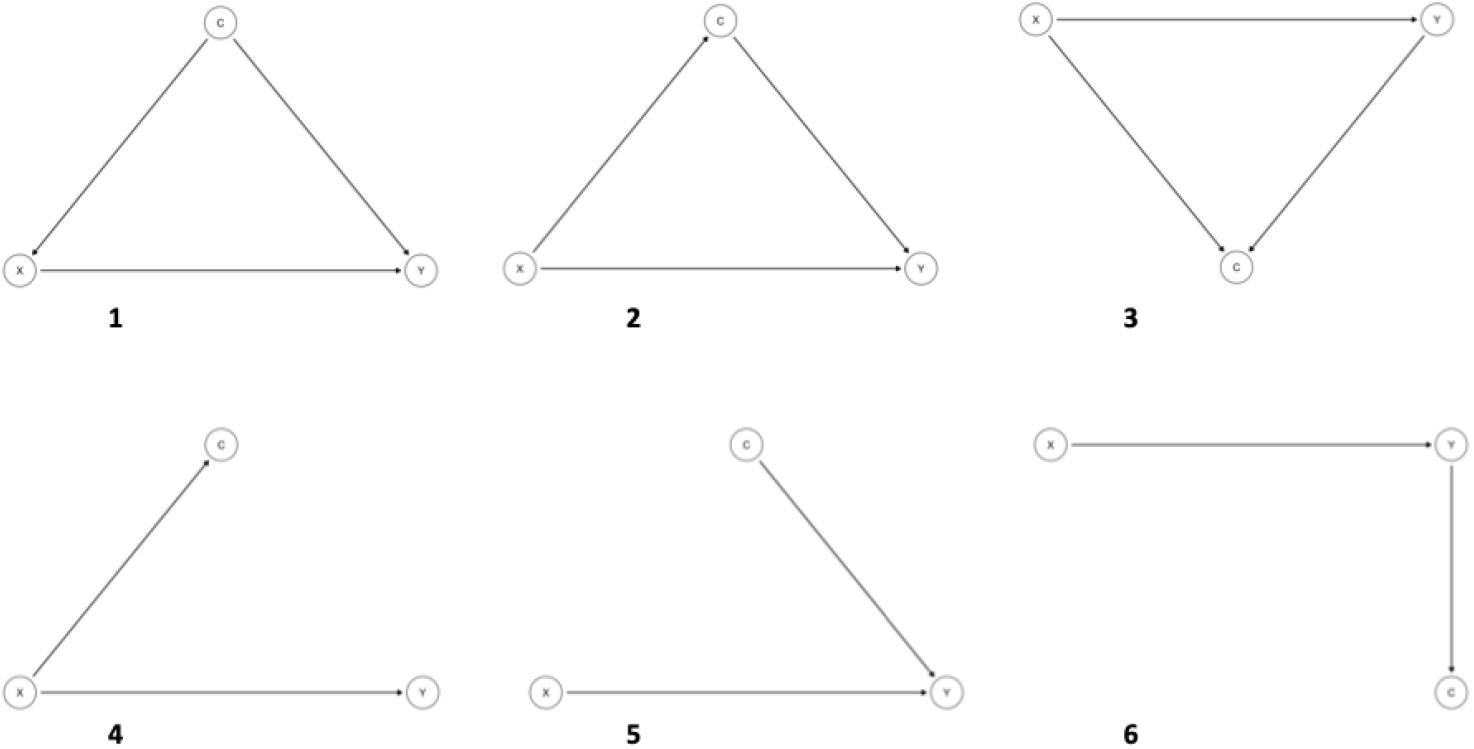
Depicting causal relation between exposure (X), index disease (Y), and comorbidity (C) in selected common scenarios using Directed Acyclic Graphs.

The simulation for the causal structure depicted in Figure 1 panel 1 included the following steps:

1. Simulate binary covariate, C, with a probability of pc.
2. Simulate a binary exposure variable, X (0, 1), where the probability of X, px, was assumed to be a function of C, P(X = 1|C).
3. Simulate a response Y, where Y was assumed to be a function of X and C.

∘ The response for the log binomial model estimating RR was simulated by: P(Y = 1|X, C) = exp(log(0.2) + log(2)*x + log(2)*c)
∘ The response for the logistic regression model estimating OR was simulated by: P(Y = 1|X, C) = 1 / (1 + exp(-(log(0.4) + log(2)*x + log(2)*c)))
∘ The response for the linear regression mode estimating mean difference was simulated by E(Y|X, C) = N~(2, 2) + N~(2*x, 2) + N~(2*c, 2)
4. Run the following models:

∘ logistic regression model estimating effect of X on Y, with and without adjusting for C. Exponentiate coefficient of X to obtain OR.
∘ log binomial regression model estimating effect of X on Y, with and without adjusting for C. Exponentiate coefficient of X to obtain RR.
∘ least square regression model estimating effect of X on Y, with and without adjusting for C.
5. Repeat steps 1-4, 5,000 times.

The point estimate for each effect measure was the median of the estimates from the 5,000 bootstraps and the lower and upper limits of the 95%CI corresponded to the 2.5^th^ and 97.5^th^ percentile of these 5,000 estimates. Given the true effect estimate for OR, RR, and least square mean difference was 2, the coefficient of x from each model was evaluated based on the relative bias (B1 −2)/2.

## RESULTS

### Anticipated effect of conditioning on comorbidity and simulation results

Table 1 presents simulation results. For each of these causal diagrams the crude and C adjusted estimated association between X and Y along with their bootstrap based 95%CI intervals were summarized on for OR, RR, and mean difference. The relative bias of the point estimate was reported as percentage.

**Table 1.** Association between exposure (X) and index disease (Y) in crude and comorbidity (C) adjusted models for different causal scenarios.

| DAG | Model | Odds ratio |  | Risk ratio |  | Least square mean difference |  |
| --- | --- | --- | --- | --- | --- | --- | --- |
|  |  | OR [95%CI] | Bias | RR [95%CI] | Bias | LSMD [95%CI] | Bias |
| 1 | Crude | 2.2 (1.71, 2.85) | 10.20% | 2.24 (1.93, 2.64) | 12.20% | 2.34 (1.89, 2.79) | 17.10% |
|  | Adjusted | 2 (1.55, 2.6) | -0.20% | 2 (1.73, 2.35) | 0.20% | 2 (1.57, 2.44) | 0% |
| 2 | Crude | 2 (1.5, 2.69) | 0.10% | 2 (1.69, 2.38) | 0.10% | 2 (1.54, 2.45) | 0% |
|  | Adjusted | 1.73 (1.27, 2.35) | -13.70% | 1.72 (1.45, 2.05) | -14.20% | 1.4 (0.93, 1.86) | -29.90% |
| 3 | Crude | 2.01 (1.55, 2.64) | 0.30% | 2 (1.71, 2.38) | -0.10% | 2 (1.65, 2.34) | 0% |
|  | Adjusted | 1.81 (1.38, 2.39) | -9.70% | 1.83 (1.55, 2.19) | -8.70% | 1 (0.71, 1.29) | -49.90% |
| 4 | Crude | 2 (1.56, 2.61) | 0% | 2 (1.69, 2.38) | 0% | 2 (1.65, 2.36) | -0.10% |
|  | Adjusted | 2 (1.55, 2.62) | 0% | 2 (1.69, 2.38) | 0% | 2 (1.65, 2.36) | 0% |
| 5 | Crude | 1.95 (1.51, 2.51) | -2.50% | 2 (1.72, 2.34) | 0.10% | 2 (1.55, 2.45) | -0.10% |
|  | Adjusted | 2 (1.54, 2.61) | 0.20% | 2 (1.75, 2.32) | 0.20% | 2 (1.57, 2.43) | 0% |
| 6 | Crude | 2 (1.54, 2.6) | 0.10% | 2 (1.78, 2.26) | 0% | 2 (1.63, 2.37) | 0% |
|  | Adjusted | 2 (1.55, 2.61) | 0.20% | 1.9 (1.7, 2.15) | -4.80% | 1.36 (1.07, 1.66) | -32.20% |

### DAG 1: Comorbidity as a common cause of X and Y

Real-world example: Obesity (C) in a study of the effects of type 2 diabetes mellitus (X) on ischemic heart disease (Y).

In this scenario, comorbidity is the common cause of exposure and outcome, fulfilling the definition of a confounder. Using DAGs, we observe the open back door path from X to C to Y, resulting in bias. Conditioning on comorbidity is warranted and failure to condition on comorbidity in this case will result in confounding of the effect estimate of X on Y. The crude estimate was biased for all three models, and the adjusted estimates remained unbiased^21^ Recommended strategy: adjusting for comorbidity.

### DAG 2: Comorbidity as a mediator of the effects of X on Y

Real-world example: hyperlipidemia (C) in a study of the effects of sedentary lifestyle (X) on ischemic heart disease (Y).

In this case, the path from X to C to Y is causal. C is a mediator on the path reflecting the indirect effect of X to Y. Therefore, conditioning on C resulted in bias in the estimate of the total effect of X on Y whereas the unconditional effect remained unbiased.

Recommended strategy: not adjusting for comorbidity.

### DAG 3: Comorbidity as a collider

Real-world example: injury (C) in a study of the effects of alcohol use (X) on depression (Y)

When the comorbidity is caused by both exposure and index disease, the path connecting exposure and index disease through comorbidity is closed and the comorbidity serves as a collider on that path. Here, the crude estimate provided unbiased effect estimates, and adjusting for C resulted in bias (collider stratification bias) in the estimates^2,4^. This bias is present regardless of the analytical model used, and its magnitude depends on the strength of the association between X and C; and Y and C.

Recommended strategy: not adjusting for comorbidity.

### DAG 4: Comorbidity as a consequence of X, but not a risk factor for Y

Real world example: liver failure (C) in a study of the effects of alcohol use (X) on dementia (Y).

In this case either conditioning on C or not will not bias the estimate of the effect of E on Y. However, even though both approaches will result in unbiased estimates, the estimated effects have a different causal interpretation and might be different. Scenario 4, both crude and C-adjusted models provided unbiased estimates of OR, RR and LSMD^3^.

Recommended strategy: both adjusting or not adjusting will not result in bias.

### DAG 5: Comorbidity as a risk factor of Y, but independent of X

Real-world example: alcohol abuse (C) in a study of the effects of air pollution (X) on hepatic cancer (Y).

When comorbidity is a risk factor for the disease but is not associated with the exposure, adjusting for comorbidity does not introduce bias. RR and LSMD are unbiased in both crude and adjusted models. For the OR, the results for the adjusted and crude models are slightly different, nevertheless none are biased (assuming model specifications are correct)^3^. However, because of the non-collapsibility of nonlinear models (e.g. logistic regression), the conditional and marginal effects of X on Y are different, and the difference in the estimates between the crude and adjusted models reflects the difference in the underlying effects being estimated, and not necessarily possible bias^24–26^. The choice of the model should be guided by clear definition of the underlying effect of interest.

Recommended strategy: both adjusting or not adjusting for comorbidity will not result in bias.

### DAG 6: Comorbidity as a consequence of Y, but not directly associated with X

Real world example: opportunistic infection (C) in a study of the effects of syringe service program (X) on risk of HIV infection (Y).

When the disease is a risk factor for comorbidity, the crude estimates are unbiased. However, under the assumption that X has an effect on Y, conditioning on comorbidity in the linear models (e.g., RR and LSMD) leads to biased estimates while the OR from the adjusted model remained unbiased^3^. None of the models will be biased by adjusting for C if the effect of X on Y is null.

Recommended strategy: both adjusting or not adjusting for comorbidity will not result in bias in OR. Not adjusting in log binomial and least squares regression models.

## DISCUSSION

This study provides a detailed exploration of the potential roles of comorbidity in the association between exposure and index disease, highlighting the diverse causal relationships that can exist between these variables. Accounting for the valid causal structure has extensive implications for causal inference in etiologic research. By systematically examining common causal structures and their effects on bias, this work addresses a critical but underexplored issue: the appropriate handling of comorbidity in causal research.

Our findings confirm that the role of comorbidity in the exposure-outcome relationship determines the impact of conditioning. When comorbidity act as a confounder, conditioning effectively mitigate confounding bias across all the effect measures (e.g., odds ratio [OR], relative risk [RR], and least-squares mean difference [LSMD]). Conversely, when comorbidity serves as a mediator or collider, conditioning introduces bias into the total effect estimate of the exposure on the outcome. This highlights the critical need to avoid inappropriate adjustments.

When comorbidity is either a cause or consequence of the exposure but has no direct connection to the outcome (except through the exposure), adjusting for comorbidity does not affect the estimated effect. However, if comorbidity is a consequence of the outcome, adjustment introduces bias in log-binomial and linear regression models, whereas such adjustment does not bias estimates in logistic regression models. Furthermore, adjusting for a risk factor of the outcome does not result in bias. However, in nonlinear models like logistic regression, the distinction between marginal and conditional effects due to the non-collapsibility of effect measures emphasizes the importance of aligning analytical strategies with study objectives, which extends beyond merely minimizing bias. In such cases marginal effects provide better insight about the effect of the exposure on the outcome on the population level, whereas, the conditional effect could be more relevant for inference on an individual level.

Although our results demonstrate that inappropriate handling of comorbidity in the analyses could lead to bias, it is important to note that the direction and magnitude of such bias depend on the distribution and prevalence of the exposure, outcome, comorbidity, and the nature of the associations among these variables. Furthermore, while our examples focused on a single comorbidity at a time, in practice, multiple comorbid conditions may be associated with the index disease. The relation of these comorbidities with the study exposure and the index disease could be different, and each should be handled according to underlying assumptions and their causal relations. Importantly, in our study, we assumed no measurement error, no selection bias, and no model misspecification. In real-life setting, and in presence of one or more of these issues, the impact of adjusting for comorbidity on the overall bias might be different than the impact of such adjustment in isolation and absence of any other source of bias.

The simulation results corroborate these theoretical insights, illustrating how the causal role of comorbidity influences bias under various analytical strategies, emphasizing the importance of explicit causal assumptions and appropriate model selection to ensure valid inferences. By examining several common causal structures between exposure, index disease and its comorbidity, this study provides practical guidance for refining study design and analytical methods. While no universal approach exists for handling comorbidity, integrating methodological rigor with clinical insight – essential to decide which of the causal structures is most applicable in the study setting - enhances the validity and applicability of etiologic research.

## Data Availability

All data produced in the present study are available upon reasonable request to the authors.

